# Cardiovascular Risk Factors in Nurses at a Teaching Hospital

**DOI:** 10.1101/2024.09.10.24313412

**Authors:** LK Fernandes, GBNC Rangel, RL Martins, DF Martin, MF Godoy

## Abstract

**Introduction:** Cardiovascular diseases [CVDs] are responsible for a large number of deaths, sick leave, and early retirements, especially among health professionals, who are constantly exposed to physical and emotional stress.

**Objective:** To identify the prevalence of modifiable and non-modifiable risk factors related to CVDs in the nursing team of a tertiary hospital in the interior of the state of São Paulo, Brazil.

**Methods:** This is a prospective-analytical epidemiological study, of a quantitative nature, conducted on 226 employees, in two stages: collection of self-declared data and measurement of anthropometric data.

**Results:** Risk factors such as family history of CVD (82.7%), alcoholism (57.1%), sedentary lifestyle (49.1%), insufficient sleep time (27.9%), high consumption of processed meals (98.2%) and sugary beverages (81.7%) were observed, among others. From anthropometric data, it was identified that 69.4% of participants were overweight (38.7%) or obese (30.6%) - evidenced by high waist circumference (48.6%) and a high waist-to-hip ratio (64.0%) - in addition to 17.1% who had blood pressure levels indicative of arterial hypertension. The majority reported not having been diagnosed with CVDs (76.1%), however, among those who did, more than half (53.7%) did not treat them properly.

**Conclusion:** It was observed that the prevalence of CVDs is not irrelevant in the group evaluated and that there are risk factors indicating a tendency for participants to develop new CVDs or aggravate existing ones - due to the presence of harmful habits associated with an unhealthy lifestyle and lack of adherence to treatments.

## INTRODUCTION

According to World Health Organization data from 2021, cardiovascular diseases [CVD] are the leading causes of death worldwide, resulting in the loss of 17.9 million lives per year, which is equivalent to 32% of global deaths. Most of these occurrences (85%) are due to heart attacks and strokes. One third of them affect people under the age of 70^1^. In a study conducted in the city of São José do Rio Preto in 2013, it was identified that CVDs were responsible for a rate of 195.9 deaths for every 100,000 inhabitants, which is equivalent to 31.7% of deaths^2^. Therefore, identifying populations at risk and mitigating modifiable factors, as well as promoting appropriate treatment can help prevent avoidable losses.

Among CVDs, the most prominent are coronary artery disease [CAD], acute myocardial infarction [AMI], heart failure [HF], high blood pressure [BP] and cerebrovascular accident [CVA]. In this context, they are influenced by risk factors that can be classified as modifiable and non-modifiable. Modifiable factors include hyperglycemia, hyperlipidemia, obesity, sedentary lifestyle, smoking, alcoholism and poor diet; non-modifiable factors include biological sex, age, ethnicity, family history, among others.

Hyperglycemia, particularly in individuals with diabetes, is a significant modifiable risk factor for CVD. Elevated blood glucose levels contribute to the development of atherosclerosis by promoting endothelial dysfunction and inflammatory processes. A study by Sun *et al*. demonstrated that individuals with diabetes have a two-fold increased risk of cardiovascular mortality compared to non-diabetics, emphasizing the importance of glycemic control as a preventive measure^3^.

Dyslipidemia, particularly elevated cholesterol and triglycerides, is another critical factor. Hypercholesterolemia has been linked to atherosclerosis, leading to CAD and CVA. Navar *et al*. reported that reductions in LDL cholesterol through statin therapy reduced the incidence of cardiovascular events, underlining the efficacy of lipid management^4^. Furthermore, hypertriglyceridemia has been identified as an independent risk factor for cardiovascular disease, as discussed by Du et al.^5^.

Obesity and a sedentary lifestyle are closely linked factors that greatly increase the risk of CVD. The accumulation of adipose tissue, particularly visceral fat, leads to metabolic disturbances such as insulin resistance, dyslipidemia, and hypertension, all of which contribute to atherosclerosis and cardiovascular events. The American Heart Association highlights that obesity, often accompanied by other risk factors like hypertension and dyslipidemia, significantly raises the risk of heart disease. Lavie *et al*. emphasized that both body weight management and increased physical activity led to significant reductions in cardiovascular mortality^6^. Similarly, the harmful effects of excessive alcohol consumption and smoking are well-documented. Studies such as those by Shield et al.^7^ and Gaiha et al.^8^ have shown their contributions to cardiovascular disease progression through oxidative stress and vascular inflammation.

In this context, CVDs remain a leading cause of morbidity and mortality among healthcare professionals, particularly nursing workers. The high physical and emotional demands of nursing, combined with irregular work hours, stress, and lifestyle factors, contribute significantly to CVD risk. Epidemiological studies have consistently reported a high prevalence of modifiable risk factors in this population. Moraes *et al*. found that nursing students exhibited high rates of alcohol consumption (56.4%), poor dietary habits (49.5%), and stress linked to professional training^9^. Similarly, Duarte-Clíments *et al*. highlighted the burden of sedentary behavior and lack of awareness regarding cardiovascular risk factors among university students in Brazil, suggesting an early onset of unhealthy habits that persist into professional life^10^. More recently, Pereira *et al*. identified a 21.8% prevalence of hypertension and 25.9% of prehypertension among Brazilian healthcare workers, with night shift work, obesity, and long working hours being key contributing factors^11^. Ferreira *et al*. further expanded on these findings, demonstrating that 75.9% of nursing professionals exhibited increased waist circumference, 43.8% were overweight, and 20.4% had diagnosed hypertension, reinforcing the critical need for workplace interventions targeting cardiovascular risk reduction^12^.

The present study aimed to obtain data on lifestyle habits, history of CVDs and current clinical status of members of the nursing team of a large teaching hospital in the interior of the state of São Paulo; as well as to identify possible modifiable factors and suggest practices for preventing CVDs.

## METHODS

This is a prospective, analytical, and quantitative epidemiological study conducted with nurses from a tertiary hospital in the interior of the state of São Paulo, Brazil (SP). This research received authorization from the nursing department management, was submitted for review by the institution’s Research Ethics Committee and obtained approval under code 5.671.736. All participants or their legal representatives were duly informed about the procedures performed and voluntarily agreed to the Informed Consent Form, according to resolution No. 466/12 of the National Health Council. The data obtained were tabulated in Excel v2308 and analyzed in StatsDirect 3.0. Normality of continuous variables was assessed using the Shapiro–Wilk test. Categorical variables were compared using the Chi-square test (χ^2^). A 95% confidence interval (CI95%) was adopted, and statistical significance was considered at a level of α = 0.05. Additionally, a multivariate logistic regression model was applied to identify independent associated variables of elevated blood pressure, considering age, sex, work hours, obesity, abdominal circumference, waist-to-hip ratio, and heart rate. The results were summarized through descriptive statistics and presented in tables. The research was divided into two phases: (1) collection of self-reported data and (2) measurement of anthropometric data. Individuals under the age of 18 were not included in the study.

In the first stage, potential participants were approached electronically and invited to participate in the study. After accepting the informed consent form, the nurses filled out an electronic form in which data on individual characteristics (sex, age, race, marital status, number of children, city of residence, occupational function); work (length of service at the institution, length of service in the health area, work shift, and number of hours worked); personal history of CVD (diabetes, hypertension, AMI, CAD, varicose veins, hypercholesterolemia, hypertriglyceridemia); and modifiable cardiovascular risk factors (smoking, alcohol consumption, sleep, mental health, sports, and eating habits) were collected. The questionnaire used is found in Appendix 1.

In the second stage, an active search was conducted for some of the participants during working hours, at a convenient time indicated by the sector head, respecting their routines and obligations. At this point, anthropometric data were measured: weight, height, abdominal circumference [AC], hip circumference [HC], heart rate [HR], systolic blood pressure [SBP], and diastolic blood pressure [DBP]. From these data, BMI, waist-hip ratio [WHR], and mean arterial pressure [MAP] were calculated.

## RESULTS

In the first stage, 226 nurses provided self-reported data, of which 111 were approached in the second stage and anthropometric data were measured. Regarding gender, a physiopathologically relevant finding was that 90.3% of the sample consisted of females. The Chi-square test indicated a statistically significant difference for this variable (χ^2^ = 146.6; P < 0.0001). In terms of age, 175 assessments involved individuals younger than 40 years (20 to 39 years), while 51 participants were 40 years or older. The Chi-square test also indicated a statistically significant difference for this variable (χ^2^ = 68.1; P < 0.0001). Most of the interviewees reside either in the city where the study was conducted (73.9%) or in cities within the same Regional Health Department. Table 1 provides a detailed description of the study population profile.

**Table 1.**
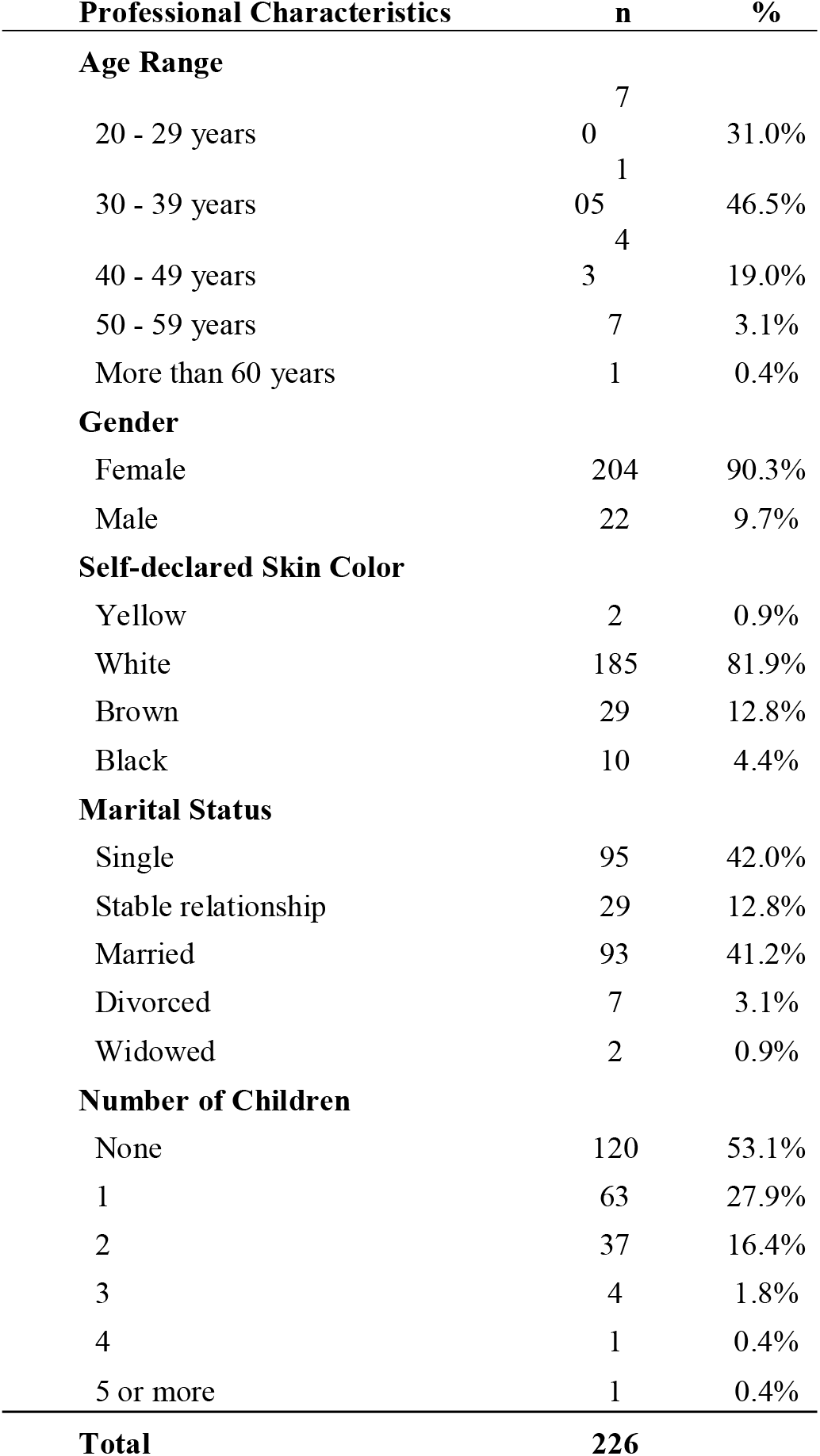
Profile of the study population.

An analysis of professionals’ employment data reveals that 51.8% of participants have less than 10 years of experience in the health sector, with the majority (62.8%) having worked for less than 5 years at the evaluated institution. Regarding work shifts, 65.3% typically work a single shift, with a 6-hour workday being the most common (42.0%).

Concerning non-modifiable cardiovascular risk factors, 82.7% reported having a family member (parents, siblings, uncles, children) with CVD. Specifically, 49.2% of employees’ fathers and 50.3% of their mothers were reported to have CVD. The most prevalent in family histories included high BP (77.0%), hypercholesterolemia (43.9%), AMI (42.8%), CVA (39.0%), and hypertriglyceridemia (39.0%).

Table 2 presents the prevalence of self-reported CDVs. Among respondents aware of their CVD condition, 53.7% were not undergoing treatment (highlighting that 25% of diabetics also do not treat it). Of those receiving treatment, 80.0% used private health insurance, while 16.0% utilized the Sistema Único de Saúde [SUS] at their workplace institution.

**Table 2.**
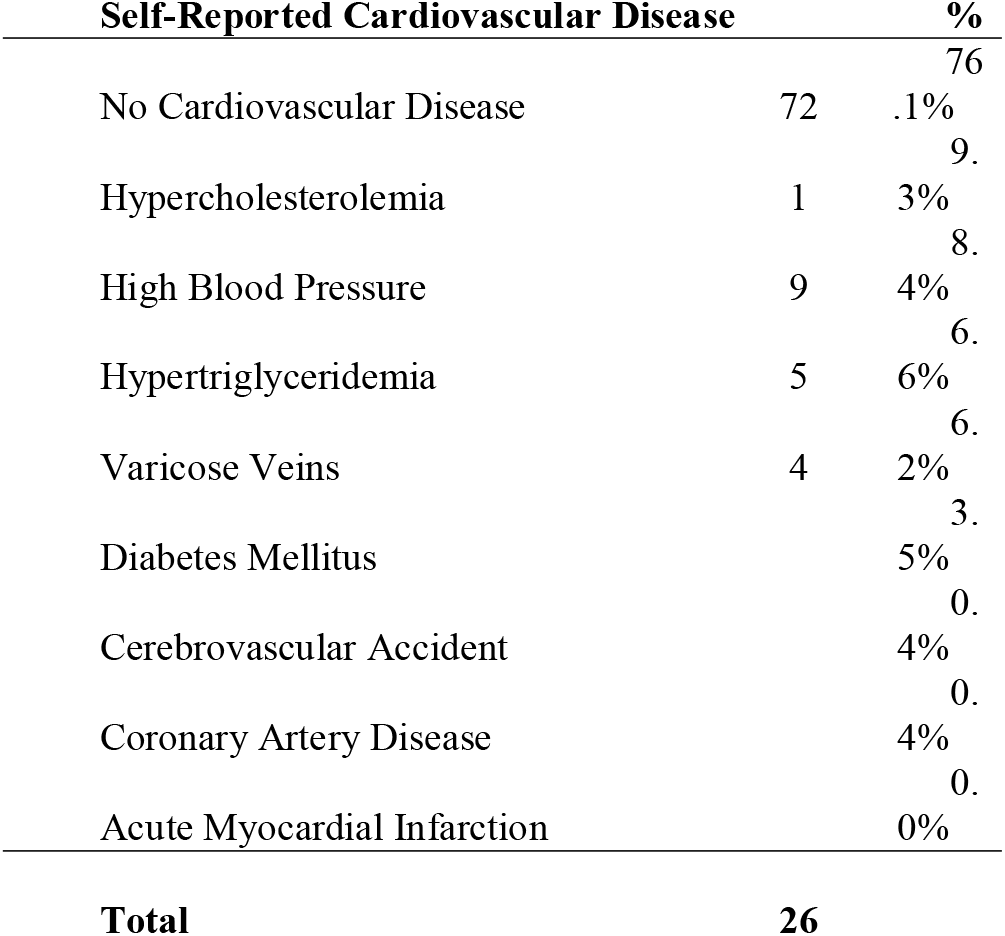
Self-reported Prevalence of Cardiovascular Diseases in Participants.

In terms of modifiable cardiovascular risk factors, the majority of participants are non-smokers (91.2%) or former smokers (5.3%). Among those who smoke or have smoked, the smoking history is low (<3 pack-years in 75%). Additionally, 42.9% of participants reported not consuming alcohol, while 54.0% reported drinking up to twice a week, and only 3.1% drank 3 to 5 times a week. A preference for beer was positively correlated with the frequency of alcohol intake (100% of those who drank more than twice per week). Occasional drinkers (up to twice per week) exhibited more diverse preferences, consuming beer (81.1%), wine (45.1%), and spirits (26.2%).

Still regarding lifestyle habits, the majority reported an adequate average sleep time (72.2% sleep > 6h/night), but more than a quarter of the participants (27.9%) sleep less than 6h per night. Furthermore, on a subjective mental health scale from 1 to 5, 58.0% self-rated their mental health as good (>3). For 70.8% of participants, their mental health status was explained by current work conditions.

A complete sedentary lifestyle was reported by almost half of the participants (49.1%) and 31.0% reported practicing less than 150 minutes of physical activity per week, so that only 19.9% exercised adequately (>150 minutes/week). Among the prevalence of sports practiced among non-sedentary individuals, weight training was the most common modality (53.3%), followed by light walking (<5 km/h; 34.8%), fast walking (>5 km/h; 15.7%), light running (10.4%) and Pilates (9.6%).

Regarding eating habits, only 2.7% are vegetarians; and, among the participants, the majority (90.2%) ate at least 1 serving of fruits, vegetables and legumes per day. More than half (52.7%) used to eat more than 3 servings of red meat per week. The consumption of processed foods is very common: 98.2% of the participants reported eating them at least once a week, while 41.2% eat them 2 to 6 times a week and 12.4% consume them once or more per day. The consumption of sugary drinks (juices, soft drinks, others) is also recurrent: 81.4% consumed at least 1 serving/day, but only 3.5% reported drinking more than 5 servings/day. In a subjective self-assessment of current diet (1 to 5 points), the majority classified themselves as 3 (1: 5.3%; 2: 18.1%; 3: 50.4%; 4: 19.5%; 5: 6.6%).

Participants were stratified based on the anthropometric data collected in the second stage, with the results shown in Table 3. A complete description of all anthropometric data is available in Appendix 2.

**Table 3.** Risk Stratification by Anthropometric Data.

| **Modifiable Risk Factors** | **n** | **%** |
| --- | --- | --- |
| **Weight Stratification (by Body Mass Index)** |  |  |
| Underweight (< 20kg/m<sup>2</sup>) | 2 | 1.80% |
| Normal Weight (20 - 25kg/m<sup>2</sup>) | 32 | 28.83% |
| Overweight (25 - 30kg/m<sup>2</sup>) | 43 | 38.74% |
| Obesity I (30 - 35kg/m<sup>2</sup>) | 21 | 18.92% |
| Obesity II (35 - 40kg/m<sup>2</sup>) | 10 | 9.01% |
| Obesity III (> 40kg/m<sup>2</sup>) | 3 | 2.70% |
| **Abdominal Circumference** |  |  |
| Normal (<88cm for women and <102cm for men) | 57 | 51.4% |
| High (>88cm for women and >102cm for men) | 54 | 48.6% |
| **Waist-to-Hip Ratio** |  |  |
| Normal (< 0.8 for women and < 0.95 for men) | 40 | 36.0% |
| High (> 0.8 for women and > 0.95 for men) | 71 | 64.0% |
| **Systemic Blood Pressure** |  |  |
| Optimal (< 120 x 80mmHg) | 51 | 45.9% |
| Normal (< 130 x 85mmHg) | 24 | 21.6% |
| Borderline (130 - 139 x 85 - 89mmHg) | 17 | 15.3% |
| Hypertension Stage I (140 - 159 x 90 - 99mmHg) | 15 | 13.5% |
| Hypertension Stage II (160 - 179 x 100 - 109mmHg) | 4 | 3.6% |
| Hypertension Stage III (> 180 x 110mmHg) | 0 | 0.0% |
| **Total** | **111** |  |

As shown in Table 3, among the 111 participants, only 32 (28.83%) had BMI values within the normal range, whereas the expected proportion, based on a realistic estimate, was 70% (approximately 78 individuals with normal BMI). A Chi-square test was conducted to compare the observed and expected frequencies. The result was highly significant (χ^2^ = 89.60; p < 0.0001), indicating that the proportion of patients with BMI within the normal range was considerably lower than expected, even when assuming a realistic reference proportion of 70%.

Among the 111 patients evaluated, 75 had blood pressure within the normal range, while 36 (32.4%) were diagnosed with hypertension. Considering a tolerable expectation of up to 10% prevalence of hypertension in the target population (corresponding to 11 expected cases), a Chi-square test was performed to compare observed and expected frequencies. The result was highly significant (χ^2^ = 62.06; p < 0.0001), indicating that the proportion of individuals with hypertension was statistically higher than the acceptable threshold.

A multivariate logistic regression analysis was performed to identify independent predictors of elevated blood pressure (systolic ≥120 mmHg and/or diastolic ≥80 mmHg). Among the tested variables, age >40 years and obesity (BMI ≥30 kg/m^2^) remained statistically significant. For age >40 years, the odds ratio was 2.52 (95% CI: 1.02–6.24; P = 0.0448), indicating that those with hypertension have a 152% greater chance of being in the >40 group than a normotensive person belonging to this age group. Similarly, for obesity, the odds ratio was 3.22 (95% CI: 1.34–7.70; P = 0.0085), indicating that a hypertensive individual is 222% more likely to be obese than a normotensive individual is. These findings suggest that age and obesity are independent predictors of increased blood pressure in this nursing population.

## DISCUSSION

The main results of this study highlight several key areas requiring intervention: insufficient physical activity, inadequate diet, lack of sleep, and a lack of proper diagnosis and treatment for CVDs. These findings align with previous research demonstrating a high prevalence of cardiovascular risk factors among nursing professionals.

To contextualize these findings on a global scale, a systematic review by Khani *et al*. analyzed cardiovascular risk factors among nurses worldwide. Their meta-analysis identified sedentary lifestyle as the most prevalent risk factor, affecting 46.3% of nurses. Furthermore, 33.3% of nurses were overweight, and 15.3% were obese - values closely resembling those observed in this study. However, a notable difference was found in alcohol consumption, with Khani *et al*. reporting a prevalence of 24.6%, whereas lower rates were observed in our cohort.

Another significant divergence was noted in the prevalence of hypertension: while the global meta-analysis reported 11.2% of nurses with SBP ≥140 mmHg, our study revealed a slightly higher prevalence. This discrepancy may be attributed to demographic, geographic, and occupational differences in the populations studied^13^. Statistical analysis of the data from the 111 participants showed that, even when considering a 10% prevalence of hypertension as tolerable within the group, the Chi-square test revealed a significant difference (χ^2^ = 62.06; p < 0.0001). This indicates that the proportion of individuals with hypertension was statistically higher than the acceptable threshold, reinforcing the importance of early screening and intervention strategies for this population.

Recognizing the impact of geographic variability, several studies have assessed cardiovascular risk factors among healthcare professionals in Brazil. The prevalence of stage I and stage II hypertension (17.1%) observed in our cohort is consistent with findings from Pereira *et al*., who reported similar rates among healthcare workers in high-complexity services in Brazil^11^.

Additionally, Ferreira *et al*. highlighted the role of central adiposity, reporting that 75.9% of nursing professionals exhibited increased waist circumference, a well-established predictor of cardiovascular morbidity and mortality^12^. In line with these findings, our multivariate analysis showed that obesity and age over 40 years were independent variables associated with elevated blood pressure. Together, these results reinforce the need for early detection and interventions aimed at weight control and lifestyle modification in this professional group.

Lifestyle factors such as physical inactivity and poor nutrition are widely recognized as key contributors to cardiovascular risk in this population. Moraes *et al*. found that unhealthy dietary patterns and alcohol consumption were prevalent among nursing students, suggesting that these risk factors develop early during academic training and persist throughout professional life^9^. Similarly, Duarte-Clíments *et al*. demonstrated that sedentary behavior and limited awareness of CVD prevention strategies were major concerns among university students, reinforcing the need for structured cardiovascular health programs at the early stages of professional development^10^. Given that our study identified a high prevalence of physical inactivity (49.1%) and poor dietary habits (98.2% consuming ultra-processed foods), workplace initiatives addressing these modifiable risk factors could be highly beneficial.

Occupational stress remains another critical determinant of CVD among healthcare workers. The demanding nature of nursing shifts, particularly night shifts, has been associated with increased rates of hypertension, metabolic disorders, and adverse cardiovascular outcomes. Pereira *et al*. found that shift workers had a significantly higher likelihood of developing hypertension compared to daytime workers, emphasizing the need for strategies to mitigate occupational stress and promote work-life balance^11^. Ferreira *et al*. similarly highlighted the association between long working hours and increased cardiovascular risk, further reinforcing the necessity of hospital policies that support healthier working conditions^12^.

Moreover, regional studies in Brazil further corroborate these findings. Magalhães *et al*. assessed cardiovascular risk factors in 165 nursing professionals in Fortaleza, CE, reporting high rates of physical inactivity (64.9%), overweight (56.4%), and increased waist-to-hip ratio (49.7%), as well as a high prevalence of family history of hypertension (72.9%)^14^. Likewise, Ulguim *et al*. examined 45 healthcare workers in Vale do Rio Pardo, RS, identifying high rates of overweight/obesity (55.5%), elevated WHR (73.4%), borderline or high cholesterol levels (88.9%), and occupational stress (55.5%)^15^. Additionally, Ramos *et al*. conducted a cross-sectional study on 188 healthcare professionals in Volta Redonda, RJ, reporting high rates of sedentary lifestyle (83.5%), alcohol consumption (35.6%), hypertension (22.9%), obesity (20.2%), smoking (19.7%), and cholesterol levels above 200 mg/dL (10.6%)^16^. The high prevalence of elevated BMI in the present study draws attention to the risk of future cardiovascular complications and strongly underscores the need for managerial action to preserve employees’ health and ensure the proper functioning of the service.

Beyond the identification of risk factors, it is important to consider possible strategies to address them in professional practice. Strategies such as implementing health promotion programs within the hospital environment, encouraging balanced diets through institutional policies, and promoting flexible work shifts to improve sleep hygiene and reduce stress may have a positive impact. While such proposals are not the primary aim of descriptive studies, the findings can offer important insights for hospital managers and policymakers when designing preventive strategies.

This study aimed to provide a cardiovascular risk profile of the nursing staff in the evaluated institution based on standardized clinical data. However, it did not include laboratory tests, which are crucial for a more comprehensive cardiovascular risk stratification. The observed prevalence of modifiable risk factors - including smoking, alcohol consumption, sedentary behavior, and overweight/obesity - aligns with findings from previous studies. Nevertheless, these studies reported higher prevalence rates of hypertension, hypercholesterolemia, and hypertriglyceridemia compared to self-reported data in our cohort, suggesting that underdiagnosis may be a concern among the participants.

Despite these contributions, some methodological limitations must be acknowledged. First, the voluntary nature of participation may have introduced a self-selection bias, with nurses more aware or concerned about health being overrepresented. Second, the absence of laboratory tests limited the depth of cardiovascular risk stratification. Finally, the predominance of women in the sample reflects the demographic reality of the nursing profession but restricted the possibility of comparing sex-related differences in cardiovascular risk. These aspects should be considered when interpreting the results and highlight the need for future studies with broader and more balanced samples.

## CONCLUSION

This study identified a high prevalence of cardiovascular diseases and modifiable risk factors among nursing professionals, including overweight, sedentary lifestyle, poor diet, and low treatment adherence. These conditions suggest not only the current burden of cardiovascular risk but also a potential worsening of outcomes if preventive measures are not implemented. The findings highlight the need for occupational health policies and tailored prevention programs for nurses. Nonetheless, limitations such as self-selection bias, absence of laboratory data, and predominance of female participants must be acknowledged. Future research should incorporate laboratory assessments, address selection bias, and adopt broader longitudinal designs to confirm and expand these results, providing stronger evidence for preventive strategies in this population.

## Data Availability

All data produced in the present study are available upon reasonable request to the authors.

## ABBREVIATIONS

AC: abdominal circumference
AMI: acute myocardial infarction
BMI: body mass index
BP: blood pressure
CAD: coronary artery disease
CI95%: 95% confidence interval
CVA: cerebrovascular accident
CVD: cardiovascular disease
DBP: diastolic blood pressure
HbA1c: hemoglobin A1C
HC: hip circumference
HF: heart failure
HR: heart rate
LDL: low-density lipoprotein
MAP: mean arterial pressure
OR: odds ratio
SBP: systolic blood pressure
SUS: Sistema Único de Saúde
TSC: total serum cholesterol
WHR: waist-to-hip ratio

## ATTACHMENTS

## APPENDIX 1

Data collection form

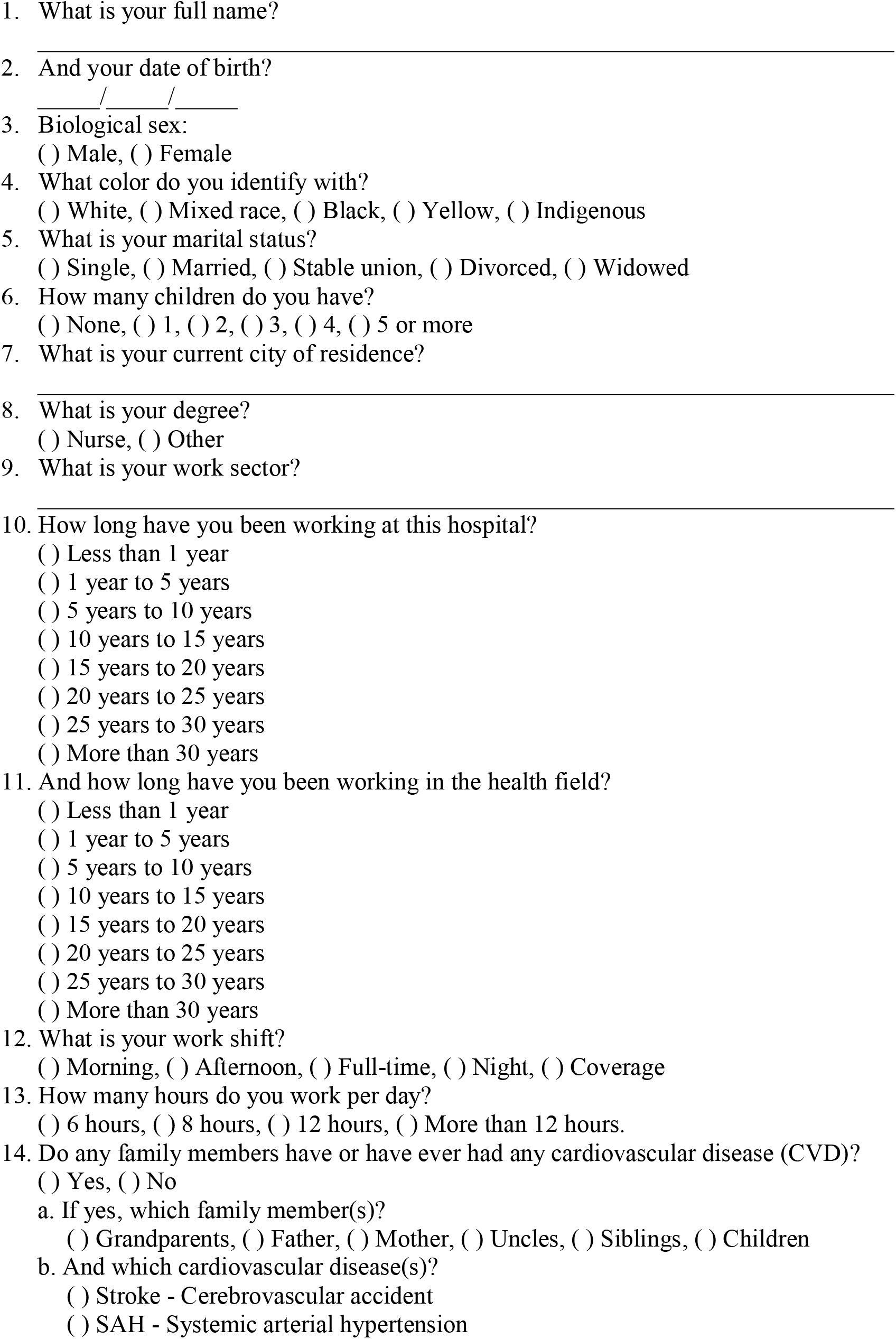
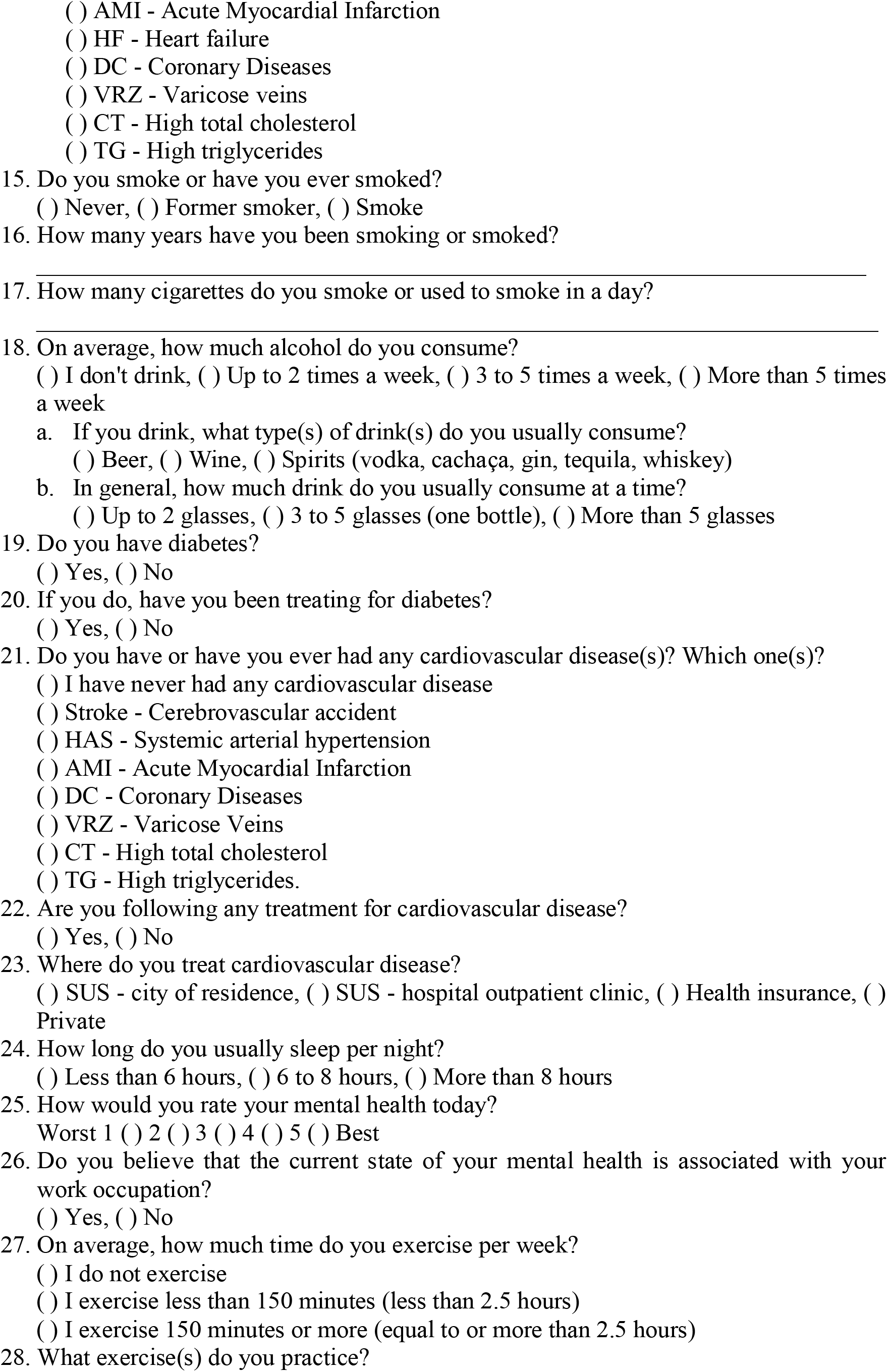
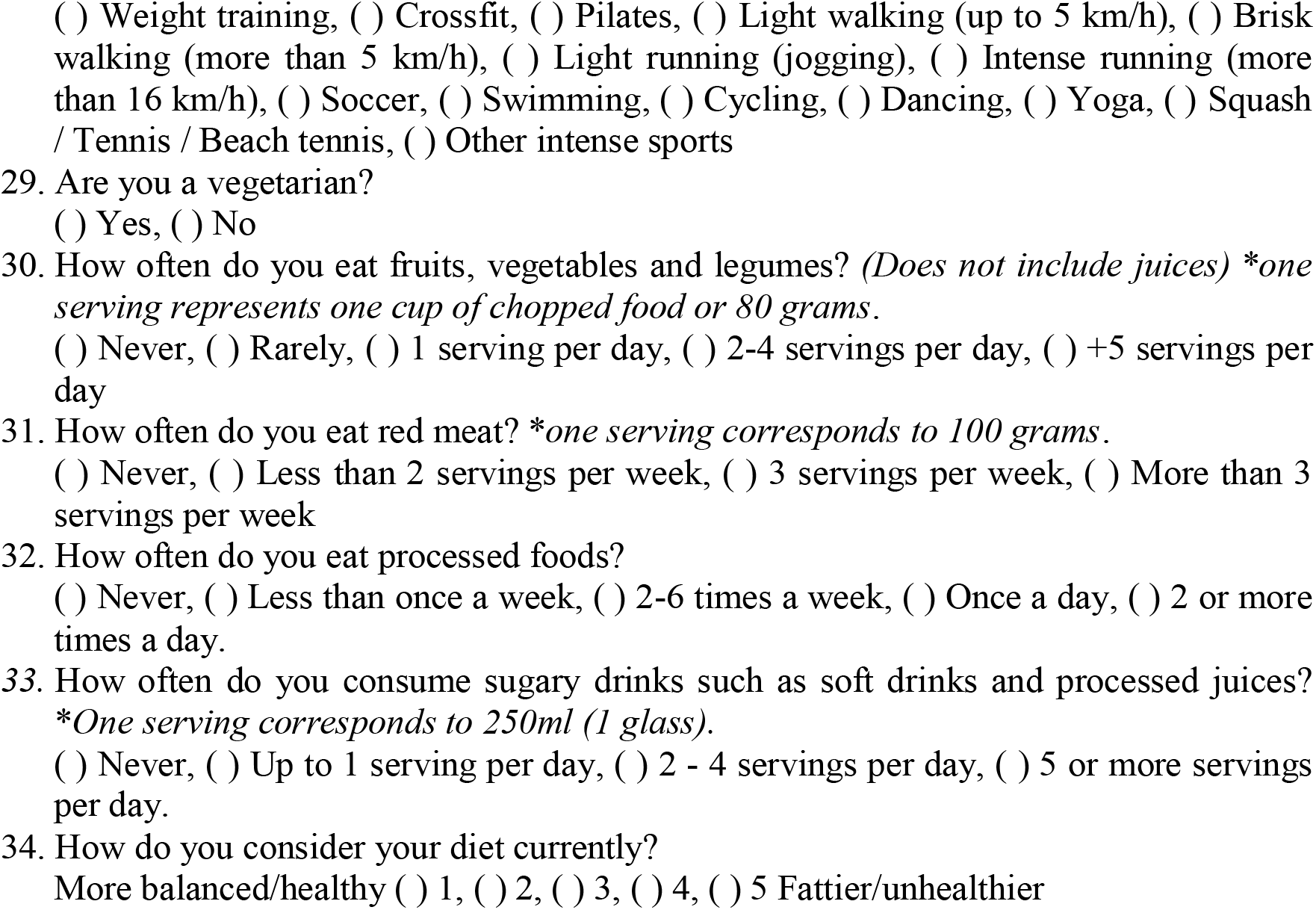

## APPENDIX 2

Distribution of Anthropometric Data

APPENDIX 2 – Distribution of Anthropometric Data
| Anthropometric Data | N | Avg | CI 95%<br>Max | CI 95%<br>Min | Max | p95 | p75 | Median | p25 | p5 | Min |
| --- | --- | --- | --- | --- | --- | --- | --- | --- | --- | --- | --- |
| Age | 221 | 34.5 | 35.6 | 33.5 | 67 | 48 | 39 | 33 | 28 | 24 | 23 |
| Age – Female | 201 | 34.5 | 35.6 | 33.4 | 67 | 49 | 39 | 33 | 28 | 24 | 23 |
| Age – Male | 20 | 35.3 | 38.4 | 32.1 | 48.0 | 46.0 | 40.5 | 35.0 | 30.0 | 24.0 | 24.0 |
| Weight | 111 | 77.3 | 80.4 | 74.2 | 114.9 | 105.4 | 91.2 | 74.0 | 64.4 | 53.3 | 43.6 |
| Weight – Female | 102 | 76.1 | 79.4 | 72.9 | 114.9 | 103.0 | 88.2 | 73.4 | 64.2 | 53.3 | 43.6 |
| Weight – Male | 9 | 91.1 | 99.3 | 82.9 | 106.7 | 106.7 | 98.4 | 91.2 | 87.2 | 70.5 | 70.5 |
| Height | 111 | 1.655 | 1.669 | 1.641 | 1.840 | 1.770 | 1.700 | 1.650 | 1.610 | 1.540 | 1.420 |
| Height - Female | 102 | 1.645 | 1.658 | 1.631 | 1.780 | 1.750 | 1.690 | 1.650 | 1.600 | 1.540 | 1.420 |
| Altura - Male | 9 | 1.771 | 1.803 | 1.739 | 1.840 | 1.840 | 1.800 | 1.750 | 1.740 | 1.730 | 1.730 |
| BMI | 111 | 28.165 | 29.209 | 27.121 | 47.475 | 39.704 | 31.846 | 27.233 | 24.583 | 20.663 | 16.914 |
| BMI - Female | 102 | 28.076 | 29.186 | 26.965 | 47.475 | 39.704 | 31.446 | 26.809 | 24.583 | 20.663 | 16.914 |
| BMI - Male | 9 | 29.178 | 32.508 | 25.849 | 35.651 | 35.651 | 32.131 | 29.442 | 27.233 | 21.759 | 21.759 |
| Abdominal Circumference | 111 | 91.4 | 93.9 | 88.9 | 134.0 | 119.0 | 101.0 | 90.5 | 81.0 | 73.0 | 71.0 |
| Abdominal Circumference - Female | 102 | 90.9 | 93.6 | 88.3 | 134.0 | 119.0 | 99.0 | 88.5 | 81.0 | 73.0 | 71.0 |
| Abdominal Circumference - Male | 9 | 97.1 | 105.1 | 89.1 | 110.0 | 110.0 | 104.0 | 98.0 | 91.0 | 76.0 | 76.0 |
| Waist Circumference | 111 | 108.4 | 110.6 | 106.3 | 140.0 | 130.0 | 116.0 | 107.0 | 100.0 | 92.0 | 86.0 |
| Waist Circumference - Female | 102 | 108.6 | 110.9 | 106.3 | 140.0 | 130.0 | 116.0 | 107.3 | 100.0 | 92.0 | 86.0 |
| Waist Circumference - Male | 9 | 106.9 | 113.7 | 100.1 | 120.0 | 120.0 | 113.0 | 105.0 | 100.0 | 95.0 | 95.0 |
| WHR | 111 | 0.842 | 0.854 | 0.829 | 1.047 | 0.948 | 0.884 | 0.838 | 0.796 | 0.748 | 0.674 |
| WHR - Female | 102 | 0.836 | 0.848 | 0.823 | 1.047 | 0.938 | 0.879 | 0.836 | 0.793 | 0.748 | 0.674 |
| WHR - Male | 9 | 0.908 | 0.948 | 0.868 | 0.982 | 0.982 | 0.933 | 0.910 | 0.894 | 0.800 | 0.800 |
| SBP | 111 | 121.8 | 124.6 | 119.1 | 165.0 | 147.0 | 131.0 | 120.0 | 111.0 | 103.0 | 93.0 |
| SBP - Female | 102 | 121.2 | 124.1 | 118.3 | 165.0 | 147.0 | 129.0 | 118.0 | 111.0 | 103.0 | 93.0 |
| SBP - Male | 9 | 129.1 | 135.3 | 122.9 | 144.0 | 144.0 | 135.0 | 126.0 | 125.0 | 119.0 | 119.0 |
| DBP | 111 | 76.3 | 78.2 | 74.3 | 103.0 | 94.0 | 83.0 | 76.0 | 69.0 | 59.0 | 52.0 |
| DBP - Female | 102 | 76.3 | 78.4 | 74.2 | 103.0 | 94.0 | 83.0 | 76.0 | 69.0 | 59.0 | 52.0 |
| DBP - Male | 9 | 75.8 | 79.9 | 71.6 | 82.0 | 82.0 | 79.0 | 78.0 | 74.0 | 67.0 | 67.0 |
| MAP | 111 | 91.5 | 93.5 | 89.4 | 123.7 | 111.0 | 99.0 | 90.3 | 83.7 | 74.3 | 68.3 |
| MAP - Female | 102 | 91.3 | 93.5 | 89.1 | 123.7 | 111.0 | 99.0 | 89.7 | 83.0 | 74.3 | 68.3 |
| MAP - Male | 9 | 93.6 | 97.6 | 89.5 | 100.0 | 100.0 | 96.3 | 94.3 | 89.0 | 85.0 | 85.0 |
| HR | 111 | 80.7 | 82.7 | 78.6 | 109.0 | 104.0 | 86.0 | 81.0 | 73.0 | 64.0 | 61.0 |
| HR - Female | 102 | 80.6 | 82.7 | 78.5 | 109.0 | 104.0 | 86.0 | 80.5 | 73.0 | 65.0 | 61.0 |
| HR - Male | 9 | 81.6 | 91.4 | 71.7 | 99.0 | 99.0 | 92.0 | 82.0 | 73.0 | 63.0 | 63.0 |

## Notes

### Competing Interest Statement

The authors have declared no competing interest.

### Funding Statement

This study did not receive any funding.

### Author Declarations

Ethics committee/IRB of Faculdade de Medicina de Sao Jose do Rio Preto (FAMERP) gave ethical approval for this work - Report number: 5.671.736.

### Summary of Updates

This version includes substantial updates to improve the scientific rigor and clarity of the manuscript. The Introduction was expanded to include recent literature and contextual data on cardiovascular risk among healthcare professionals. The Methods section now describes additional statistical analyses, including tests for normality, Chi-square comparisons, and a multivariate logistic regression model identifying independent predictors of elevated blood pressure. The Results were refined to incorporate these analyses and to better present statistically significant findings. The Discussion was rewritten for greater depth and integration with national and international data, and a dedicated Limitations paragraph was added to address methodological constraints. Minor stylistic and structural edits were also made to improve readability and coherence.

